# Tuberculosis treatment outcomes after transfer or release from incarceration: A retrospective cohort study from Brazil

**DOI:** 10.1101/2025.04.19.25325982

**Authors:** Yasmine Mabene, José Bampi, Everton Ferreira Lemos, Roberto de Oliveira, Crhistinne Gonçalves, Maria de Lourdes Delgado Alves, Maridiane Coutinho Echevarria, Julio Croda, Jason R Andrews, Yiran E Liu

## Abstract

**Background:** Tuberculosis (TB) disproportionately affects people deprived of liberty. Prior studies have shown higher TB treatment completion rates among incarcerated individuals compared to the general population. However, little is known about how incarceration-related movements such as transfers between facilities or releases to the community affect TB treatment outcomes.

**Methods:** We linked person-level incarceration data with TB notifications data from the Notifiable Disease Information System for the Brazilian state of Mato Grosso do Sul between January 2006 and December 2018. We constructed a cohort of individuals newly diagnosed with drug-susceptible TB who initiated treatment while incarcerated. We compared treatment outcomes between individuals who remained in the same carceral facility and those who were transferred to other facilities or released from incarceration during treatment. We computed the covariate-adjusted relative risk of unfavorable treatment outcomes for individuals transferred or released during treatment.

**Results:** We identified 1,274 individuals who initiated TB treatment while incarcerated. Of these individuals, 849 (66.6%) remained in the same carceral facility, 259 (20.3%) were transferred to other facilities, and 166 (13.0%) were released to the community during treatment. Among those who remained in the same carceral facility, 72.3% (614/849) were successfully treated within eight months following treatment initiation. In contrast, only 61.0% (158/259) of those who were transferred and 49.4% (82/166) of those who were released achieved TB treatment success within eight months. After adjusting for covariates, the risk of unfavorable treatment outcomes was 1.4 (95% CI: 1.2 to 1.7) times as high for individuals transferred to other facilities and 1.6 (95% CI: 1.3 to 2.0) times as high for individuals released from incarceration, compared to those who remained incarcerated in the same facility during treatment. For individuals released less than two months into treatment, the risk of unfavorable treatment outcomes was twice as high (adjusted relative risk [aRR]:2.1, 95% CI: 1.6 – 2.6).

**Conclusions:** Transfers between facilities and releases from incarceration are common and may pose barriers to TB treatment completion. Strategies for ensuring continuity of care across carceral facilities and between prison and community health systems are urgently needed to improve TB outcomes for individuals impacted by incarceration.

## Background

In 2023, 10.8 million people fell ill with tuberculosis (TB) worldwide, and 1.25 million people died from TB [1]. Persons deprived of liberty (PDL) face increased risk of TB compared to the general population due to factors such as prison overcrowding, poor ventilation, undernutrition, and limited healthcare access [2–3]. High rates of TB in prisons can also spill over into communities through prison staff, visitors, and individuals released from incarceration [4–5]. Targeted efforts to address TB in prisons are needed to reduce TB morbidity and mortality for PDL and to progress toward population-wide TB elimination.

In 2023, the United Nations High-Level Meeting established global targets aimed at ensuring a 90% treatment completion rate for all people with TB [6]. Achieving these targets is especially crucial for vulnerable populations, including PDL. Although previous studies have reported higher TB treatment completion rates for PDL compared to the general population, these rates still fall short of the UN’s 90% target [7–12]. More research is needed to identify barriers and facilitators to treatment completion among PDL to improve TB outcomes in this population.

Individuals who begin TB treatment while incarcerated may face interruptions to care following transfers between carceral facilities or release from incarceration. Research has shown this for incarcerated people living with HIV, finding that releases from jails and prisons may disrupt the HIV continuum of care [13–15]. Similar challenges have been observed among people undergoing treatment for substance use disorders, including low rates of treatment retention post-release [16–17]. Studies in the United States on TB preventive therapy for people with latent tuberculosis infection (LTBI) have found lower completion rates for individuals released during treatment, with some studies reporting rates as low as 20% [18–20]. Far less is known about how carceral movements impact treatment continuity and outcomes for individuals with active TB. Studies in Uganda and Zambia found treatment disruptions and increased rates of loss to follow up for incarcerated individuals who were transferred or released during treatment, as well as for those residing in prisons with shorter incarceration sentences [21–23]. However, there remains a significant knowledge gap regarding TB treatment outcomes and linkage to care among individuals experiencing carceral movements in other regions and settings.

In Brazil, incarceration is the leading driver of the TB epidemic, accounting for approximately 37% of new cases [24]. Brazil, which has the third largest prison population in the world, has the highest number of TB cases among incarcerated individuals [25–26]. Despite this, carceral movements for individuals diagnosed with TB in Brazil are not well documented, and the impact of carceral movements on TB treatment in Brazil remains unknown. PDL in Brazil may reside in closed prisons, semi-open prisons (where individuals leave prison during the day for work and return at night), or in police detention. Varying frequencies of movements and healthcare resources across carceral facilities may affect treatment outcomes and continuity of care. As both TB incidence and incarceration rates rise in Brazil, it is essential to investigate these potential effects to inform strategies for improving TB treatment outcomes for incarcerated individuals.

In this study, we link individual-level incarceration and TB notifications data for the Brazilian state of Mato Grosso do Sul to assess the impact of carceral movements on TB treatment outcomes. We examine the proportion of individuals experiencing transfers or releases during TB treatment and determine the relative risk of unfavorable treatment outcomes compared to individuals who remain in the same carceral facility during treatment.

## Methods

### Study design

We conducted a retrospective cohort study of individuals who were newly diagnosed with drug-sensitive TB (pulmonary and extra-pulmonary) and who initiated treatment while incarcerated in Mato Grosso do Sul state, Brazil, between January 1^st^, 2006 and June 30^th^, 2018. We examined TB treatment outcomes among individuals who were transferred to other carceral facilities and/or released from incarceration during the six-month treatment period, compared to those remaining in the same carceral facility. We included individuals who were incarcerated in closed prisons, semi-open prisons, or police detention at the time of treatment initiation. We excluded individuals with prior TB diagnoses, those with drug resistance at the time of diagnosis, and those under the age of 18. The study was approved by the institutional review boards (IRBs) of Federal University of Mato Grosso do Sul (UFMS) (#7.060.054) and Stanford University (#50466).

### Data sources

We accessed statewide individual-level data on TB notifications and treatment outcomes between January 1^st^, 2006 and June 30^th^, 2020 from *Sistema de Informação de Agravos de Notificação* (SINAN). To determine incarceration status and movements during and after TB treatment initiation, we linked SINAN with Sistema Integrado de Gestão Operacional (SIGO), a database containing movements within the carceral system (i.e., entries, transfers, and releases) in Mato Grosso do Sul from January 1^st^, 2006 to June 30^th^, 2018. To account for incomplete data on deaths during the follow-up period, we performed additional linkage with the mortality database, Sistema de Informações Sobre Mortalidade (SIM), accessed between January 1^st^, 2006 and June 30^th^, 2020.

### Data processing and linkage

For the incarceration (SIGO) and TB (SINAN) databases, where an individual could have multiple distinct entries (corresponding to multiple carceral movements, TB diagnoses, etc.), we identified records belonging to unique individuals using approximate string matching over name and mother’s name, with additional comparisons of identification numbers and dates of birth when available (Text S1, Additional File 1). Approximate string matching was performed using stringdist in R version 4.4.1 [27–28].

We linked unique individuals across the incarceration and TB databases using approximate string matching on shared identifiers (name and mother’s name), resulting in a sensitivity of 90% and specificity of 95% (Text S1: SIGO and SINAN database linkage approach, Additional File 1). We performed subsequent filtering steps to exclude false positive matches (Text S1: SINAN and SIGO cohort filtering, Additional File 1). Among matched individuals, we then searched for deaths within the mortality (SIM) database using name, mother’s name, and date of birth. Although the TB (SINAN) database includes information on deaths, we additionally matched individuals to the mortality database (SIM) to account for unreported deaths (Text S1: SIGO, SINAN, and SIM database linkage approach, Additional File 1). Exact criteria for database linkage can be found in (Figure S1, Additional File 1).

### Group assignment and outcome determination

We categorized individuals by whether they experienced carceral movements (transfers to other facilities or release from incarceration) within six months following treatment initiation. Each individual was assigned to one of three groups: 1) individuals who remained in the same carceral facility throughout the treatment period (the “stationary” group); 2) individuals who were transferred to another carceral facility at least once but were not released (the “transferred” group); and 3) individuals who were released from incarceration (the “released” group; this group may also include individuals who escaped incarceration). To avoid exposure misclassification, only movements that occurred prior to ascertainment of treatment outcomes were considered in group assignment (Text S1: Group and Outcome Ascertainment, Additional File 1). In an additional analysis, we stratified the released group into two subgroups based on the timing of release (released within or after two months following treatment initiation).

In the primary analysis, we evaluated TB treatment outcomes at eight months post treatment initiation. While treatment completion takes six months, we used the eight-month point to account for delays in reporting treatment outcomes. We additionally assessed treatment outcomes at two years post treatment initiation. We considered a treatment outcome to be positive if the individual achieved treatment success; all other outcomes were considered unfavorable treatment outcomes. We categorized treatment outcomes in our study using the SINAN guidelines as follows [29]:

1. **Treatment success:** Patients who obtained two negative sputum smears (one at any follow-up month and one at the end of treatment) or those who completed treatment without evidence of treatment failure and were discharged based on clinical and radiological criteria, in accordance with Brazilian guidelines [29].
2. **No case status update:** Patients whose case had no updates in the TB (SINAN) database following treatment initiation.
3. **No case status update following treatment discontinuation/transfer and resumption of care:** Patients who discontinued treatment or transferred health facilities and were later linked to care (resumed treatment) without further case updates in the TB (SINAN) database.
4. **Treatment discontinuation with no follow-up record:** Patients who interrupted treatment for 30 or more consecutive days and had no subsequent updates in the TB (SINAN) database.
5. **Transferred health facility with no follow-up record:** Patients who were transferred to another health facility and had no subsequent updates in the TB (SINAN) database.
6. **Drug-resistant TB (DR-TB):** Patients with confirmed resistance to any anti-tuberculosis drug after treatment initiation.
7. **Death (TB):** Patients with tuberculosis as the cause of death.
8. **Death (non-TB):** Patients with cause of death other than tuberculosis.
9. **Change of diagnosis/regimen:** Patients whose initial TB diagnosis was incorrect or patients who changed treatment regimen due to drug intolerance or toxicity

### Statistical Analysis

We used the chi-square test to compare characteristics across groups and to assess differences in the proportion of treatment success across carceral movement groups. To determine the effect of carceral movements on treatment outcomes, we performed multivariable Poisson regression, adjusting for race, age, education, alcohol use disorder, mental illness, incarceration history in the past two years (total duration and unique entries into the carceral system), and facility type at treatment initiation (closed prison, semi-open prison, police detention) [30]. We included facility type at treatment initiation as a covariate due to variation across carceral facilities in healthcare, security level, and movements. Covariates with less than 25% missingness were imputed using missForest version 1.5 in R (Table S1, Additional File 1) [31]. For covariates with greater missingness, we retained “unknown/missing” as a level in the regression. For the regression analysis, we restricted our cohort to individuals who initiated TB treatment after January 1^st^ 2007. This enabled adjustment for incarceration history in the past two years.

We conducted two sensitivity analyses. First, we excluded from the regression individuals who died from non-TB causes within the first six months following treatment initiation. This exclusion was made to mitigate potential survival-induced non-exchangeability bias, as individuals in the transferred or released group had to survive long enough to experience carceral movements. In a second sensitivity analysis, we adjusted for individual carceral facility in the regression model to account for potential differences in healthcare across facilities.

## Results

### Baseline Cohort Characteristics

After database linkage, we obtained a cohort of 1,274 individuals who initiated TB treatment while incarcerated between January 1, 2006 and June 30, 2018 (Figure 1). Our cohort primarily consisted of male individuals under the age of 30 (Table 1). Nearly half of participants were Black or mixed race (N=584, 45.8%), and the majority (N=690, 54.2%) indicated not having completed high school. Additionally, 86 (6.8%) individuals reported having alcohol use disorder, and less than 1% (9, 0.7%) were reported to have mental illness. The majority of individuals (950, 74.6%) resided in closed prisons at the time of treatment initiation. In the two years preceding treatment initiation, 1021 (80.1%) had been incarcerated for more than one year and approximately one in four individuals (333, 26.1%) had experienced multiple incarcerations.

**Figure 1:**
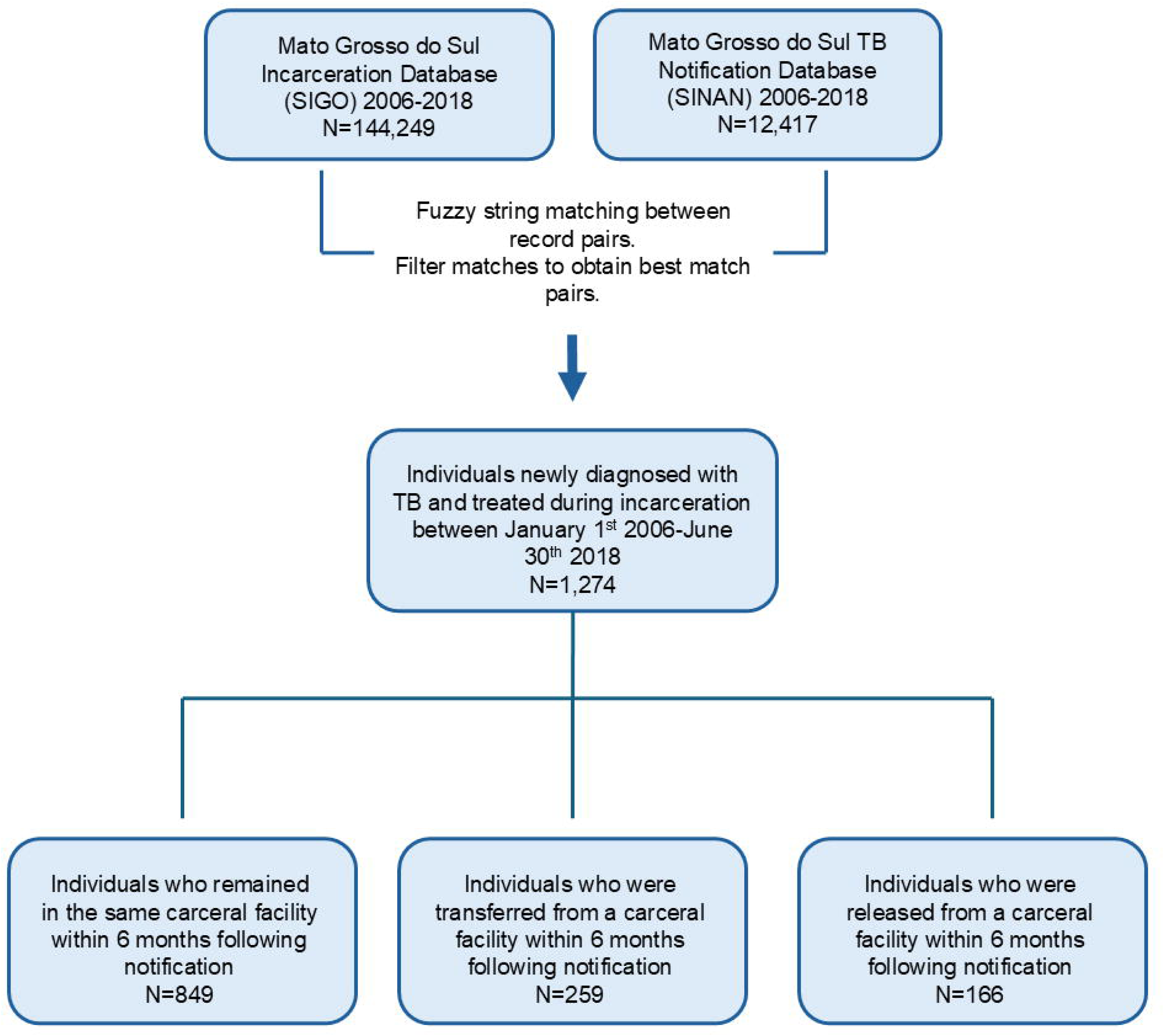
Database Linkage between SIGO and SINAN. Identification of individuals who were diagnosed with TB and initiated TB during incarceration, and classification by carceral movements during treatment. See Text S1, Additional File 1 for matching and inclusion criteria. SIGO, Sistema Integrado de Gestão Operacional (incarceration database); SINAN, Sistema de Informação de Agravos de Notificação (TB notifications database).

**Table 1.** Demographic and clinical characteristics of individuals grouped by carceral movement during TB treatment.

| Characteristics | Categories | Stationary N=849 | Transfer N=259 | Release N=166 | p-value |
| --- | --- | --- | --- | --- | --- |
| Facility type | Closed prison | 659 (77.6%) | 205 (78.5%) | 86 (51.8%) | <.001 |
|  | Semi-open prison | 100 (11.8%) | 30 (11.5%) | 58 (34.9%) |  |
|  | Police detention | 90 (10.6%) | 24 (9.2%) | 22 (13.3%) |  |
| Sex | Male | 827 (97.4%) | 254 (98.1%) | 158 (95.2%) | 0.18 |
|  | Female | 22 (2.6%) | 5 (1.9%) | 8 (4.8%) |  |
| Age | 18-29 | 419 (49.3%) | 148 (57.1%) | 76 (45.8%) | 0.14 |
|  | 30-44 | 335 (39.5%) | 94 (36.3%) | 69 (41.6%) |  |
|  | >45 | 92 (10.8%) | 17 (6.7%) | 20 (12.0%) |  |
|  | Not reported | 3 (0.4%) | 0 (0.0%) | 1 (0.6%) |  |
| Race | White | 281 (33.1%) | 82 (31.7%) | 50 (30.1%) | 0.75 |
|  | Mixed Race | 331 (39.0%) | 93 (35.9%) | 73 (44.0%) |  |
|  | Black | 59 (6.9%) | 18 (6.9%) | 10 (6.0%) |  |
|  | Asian | 12 (1.4%) | 4 (1.5%) | 1 (0.6%) |  |
|  | Indigenous | 15 (1.8%) | 8 (3.1%) | 5 (3.0%) |  |
|  | Not reported | 151 (17.8%) | 54 (20.8%) | 27 (16.3%) |  |
| Education | Did not complete high school | 453 (53.4%) | 152 (58.7%) | 85 (51.2%) | 0.08 |
|  | Completed high school or more | 28 (3.3%) | 9 (3.5%) | 12 (7.2%) |  |
|  | Not reported | 368 (43.3%) | 98 (37.8%) | 69 (41.6%) |  |
| Alcohol use disorder | No | 591 (69.6%) | 181 (69.9%) | 110 (66.3%) | 0.45 |
|  | Yes | 53 (6.2%) | 16 (6.2%) | 17 (10.2%) |  |
|  | Not reported | 205 (24.1%) | 62 (23.9%) | 39 (24.7%) |  |
| Mental illness | No | 647 (76.2%) | 202 (78.0%) | 122 (73.5%) | 0.39 |
|  | Yes | 5 (0.6%) | 1 (0.4%) | 3 (1.8%) |  |
|  | Not reported | 197<br>(23.2%) | 56 (21.6%) | 41 (24.6%) |  |
| **Total time incarcerated in past two years** | <1 year | 156<br>(18.4%) | 44 (17.0%) | 53 (31.9%) | <.001 |
|  | 1-2 years | 693<br>(81.6%) | 215<br>(83.0%) | 113<br>(68.1%) |  |
| **Number of distinct incarcerations\* in past two years** | 1 | 658<br>(77.5%) | 169<br>(65.3%) | 114<br>(68.7%) | <.001 |
|  | >1 | 191<br>(22.5%) | 90 (34.7%) | 52 (31.3%) |  |

Facility type refers to location at time of TB notification. \*Distinct incarcerations defined as periods of incarceration following time in the community. P-values are computed using chi-square test of independence with Monte Carlo simulations for categories with frequencies <5.

The majority of individuals in the cohort (849, 66.6%) remained in the same carceral facility in the six months following TB treatment initiation. Another 259 (20.3%) individuals were transferred to other carceral facilities, and 166 (13.0%) were released to the community within six months of treatment initiation (Table 1). Individuals who initiated treatment in semi-open prisons were more likely to be released to the community during treatment compared to those who initiated treatment in any other facility type (Figure S2, Additional File 1). At the time of treatment initiation, a higher proportion of individuals in the released group had spent less than one of the past two years incarcerated (31.9%), compared to individuals in the stationary (18.4%) and transferred (17.0%) groups (p value <.001) (Table 1). Lastly, the stationary group had a lower proportion of individuals with multiple prior incarcerations in the last two years (22.5%), compared to the transferred (34.7%) and released (31.3%) groups (p value <.001). The distributions of the year of TB notification and carceral facility at time of treatment initiation can be found in Tables S2-S3, Additional File 1.

### TB treatment outcomes by carceral movement group

Across the entire cohort, 854 (67.0%) of individuals successfully completed treatment within eight months following treatment initiation. Another 179 (14.0%) individuals were lost to follow up, 187 (14.7%) had no case status updates, and 38 (3.0%) died (Table 2). Among those who remained in the same carceral facility in the six months following TB notification, 614 (72.3%) had successful treatment, 84 (9.9%) were lost to follow up, 110 (13.0%) had no case status updates, and 33 (3.9%) died. Compared to those who remained in the same carceral facility, those who experienced movements during treatment had significantly lower treatment success rates: 61.0% in the transferred group and 49.4% in the released group (p-value <.001). At two years following treatment initiation, success rates remained lower for those transferred (64.9%) or released (54.2%) compared to those who remained stationary (73.7%) (p-value <.001) (Table S4, Additional File 1). Additionally, the released group was less likely to have updates to their treatment status reported in SINAN within eight months of notification, with 22.3% missing case status updates compared to only 13.0% in the stationary group (p-value <.01).

**Table 2.**
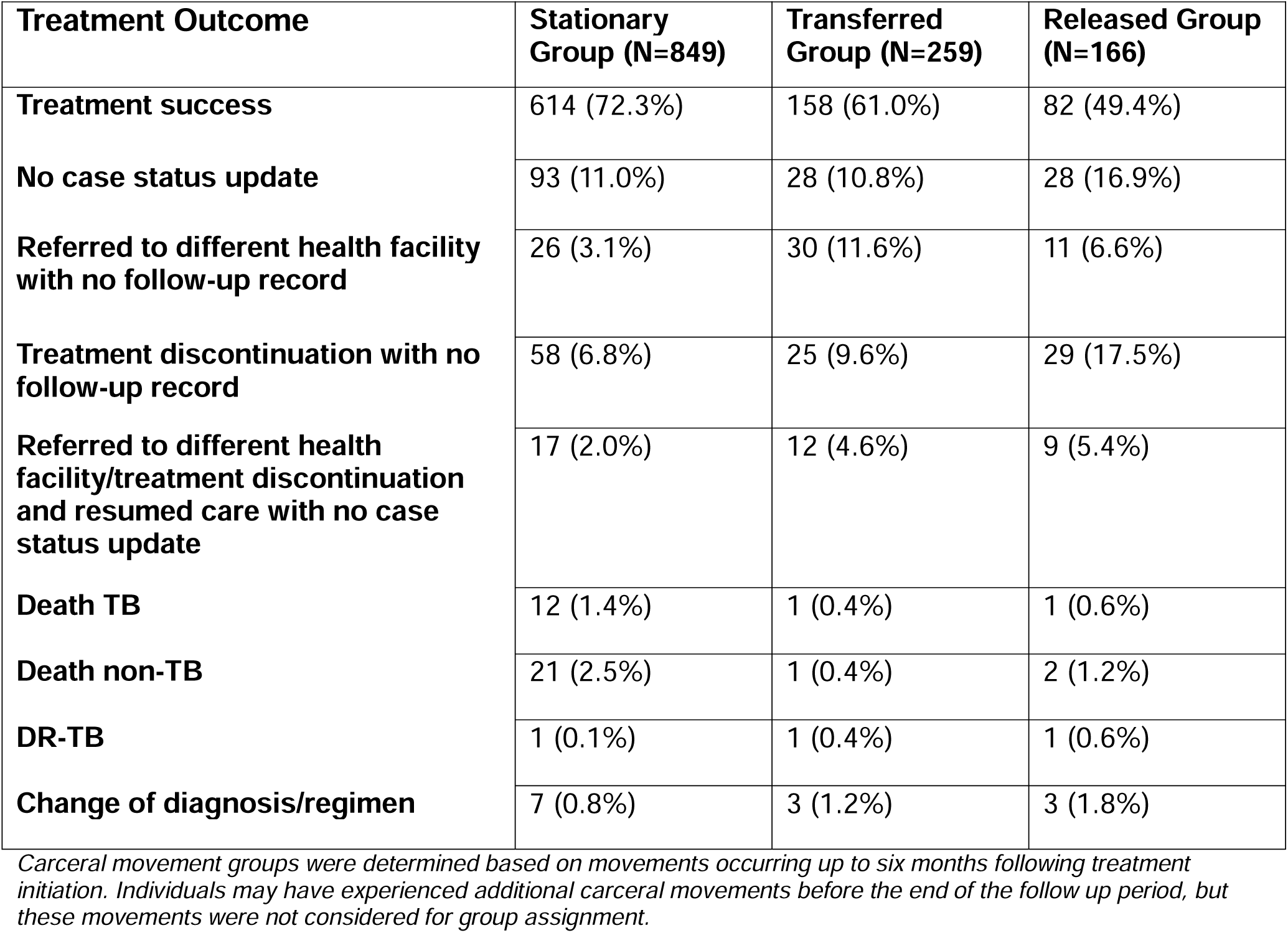
TB treatment outcomes eight months following treatment initiation.

| **Treatment Outcome** | **Stationary Group (N=849)** | **Transferred Group (N=259)** | **Released Group (N=166)** |
| --- | --- | --- | --- |
| **Treatment success** | 614 (72.3%) | 158 (61.0%) | 82 (49.4%) |
| **No case status update** | 93 (11.0%) | 28 (10.8%) | 28 (16.9%) |
| **Referred to different health facility with no follow-up record** | 26 (3.1%) | 30 (11.6%) | 11 (6.6%) |
| **Treatment discontinuation with no follow-up record** | 58 (6.8%) | 25 (9.6%) | 29 (17.5%) |
| **Referred to different health facility/treatment discontinuation and resumed care with no case status update** | 17 (2.0%) | 12 (4.6%) | 9 (5.4%) |
| **Death TB** | 12 (1.4%) | 1 (0.4%) | 1 (0.6%) |
| **Death non-TB** | 21 (2.5%) | 1 (0.4%) | 2 (1.2%) |
| **DR-TB** | 1 (0.1%) | 1 (0.4%) | 1 (0.6%) |
| **Change of diagnosis/regimen** | 7 (0.8%) | 3 (1.2%) | 3 (1.8%) |

*Carceral movement groups were determined based on movements occurring up to six months following treatment initiation. Individuals may have experienced additional carceral movements before the end of the follow up period, but these movements were not considered for group assignment.*

The proportion of individuals who were lost to follow up after being referred to a different health facility was over three times as high for individuals who were transferred to other carceral facilities (11.6%) compared to those who remained in the same location (3.1%) (p-value <.001). For individuals who were released, this proportion was more than twice as high (6.6%) (p-value = .04). Among those released from carceral facilities who were referred to another health facility or experienced treatment discontinuation within eight months of notification, 18 (36.7%) were subsequently linked to care, and 10 (20.4%) successfully completed treatment within the state over a two-year period.

### Factors associated with unfavorable treatment outcomes

After adjusting for potential confounders, transfer between carceral facilities during TB treatment was associated with a 1.4-fold (95% CI: 1.2 to 1.7) increased risk of unfavorable treatment outcomes compared to the stationary group (Figure 2). For individuals released from incarceration during treatment, the covariate-adjusted risk of unfavorable treatment outcomes was 1.6 (95% CI: 1.3 to 2.0) times as high as individuals in the stationary group. Results were similar in a sensitivity analysis excluding individuals who died from causes other than TB within six months following treatment initiation (Table S6, Additional File 1).

**Figure 2:**
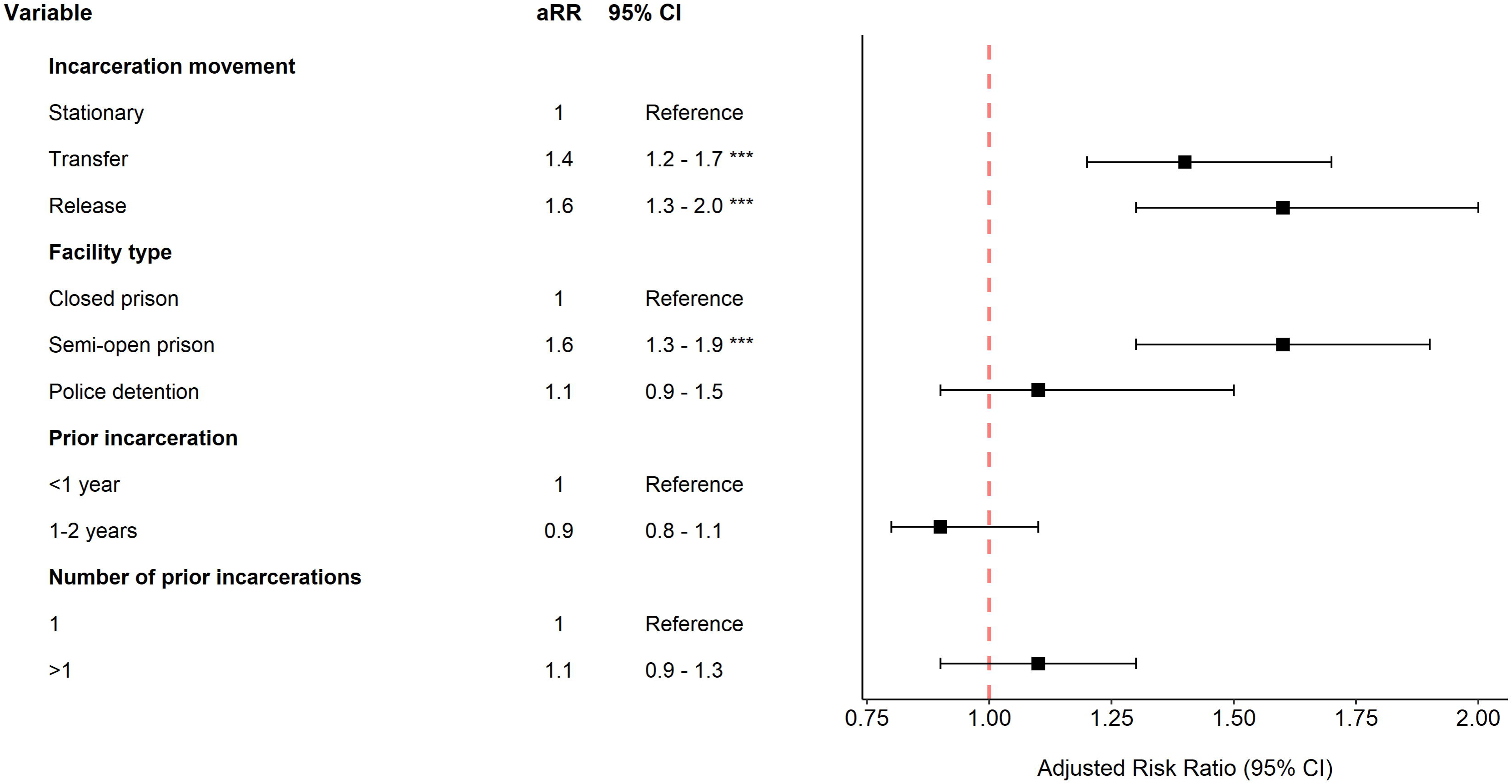
Adjusted relative risks associated with unfavorable treatment outcomes within 8 months of notification (carceral variables) Adjusted relative risks of unfavorable treatment outcomes evaluated eight months after date of notification for incarceration-related variables. Unfavorable outcomes refer to all outcomes other than treatment success. The duration of previous incarceration and number of prior arrests were recorded in the two years preceding the date of notification. Facility type refers to the facility at the time of TB notification. Adjusted relative risks for date of notification can be found in (Table S5, Additional File 1).

Beyond carceral movements, residing in a semi-open prison at the time of treatment initiation was associated with increased risk of unfavorable treatment outcomes (aRR: 1.6, 95% CI: 1.3 to 1.9) (Figure 2). However, this association did not remain after adjusting for individual carceral facility units in the regression (Table S6, Additional File 1). Additionally, higher education level was associated with a significantly reduced risk of unfavorable treatment outcomes (aRR: 0.5, 95% CI: 0.3 to 0.9) (Figure S3, Additional File 1).

### Treatment outcomes based on time of release

Among individuals in the released group, 40.4% were released less than two months after treatment initiation, during the intensive phase of TB treatment (Figure S4, Additional File 1). The treatment success rate within eight months was significantly lower among individuals released less than two months after initiating TB treatment (35.8%) compared to those who were released two or more months into treatment (58.6%) (p-value <.01) (Figure 3). Additionally, the proportion of individuals lost to follow up was 37.3% among those released less than two months after treatment initiation, compared to 24.2% among those released two months or later.

**Figure 3:**
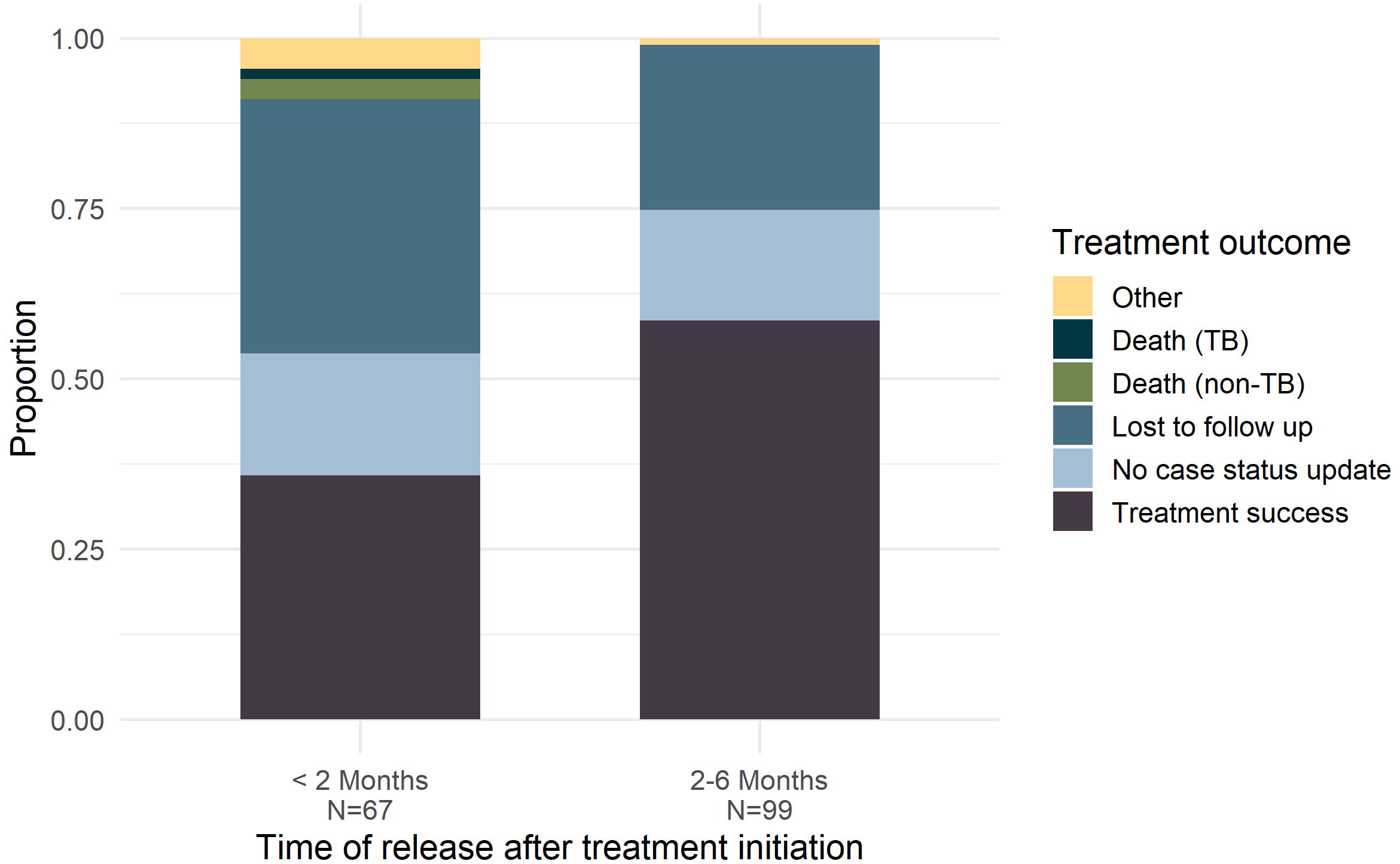
Tuberculosis treatment outcomes stratified by time of release. Bar plots reporting treatment outcomes of individuals released less than two months or two months or more from the date of notification. No case status update includes individuals without any updates to their status in the TB database following treatment initiation, as well as individuals who were transferred to new health facilities/discontinued treatment, restarted treatment, and had no subsequent updates to their case status. Other refers to individuals who experienced change of diagnosis/regimen or were diagnosed with DR-TB.

Release from incarceration within the first two months of TB treatment was associated with more than twice the risk of unfavorable treatment outcomes, compared to those who were stationary (aRR 2.1, 95%CI: 1.6 to 2.6) (Table S7, Additional File 1). For individuals released two months or more following treatment initiation, risk of unfavorable treatment outcomes was 1.3 (95% CI: 1.0 to 1.8) times that of the stationary group.

## Discussion

Timely diagnosis and treatment completion are essential to reduce morbidity and mortality among TB patients and reduce onward transmission, particularly in high-risk environments like prisons. However, in settings with limited prison healthcare resources and barriers to coordination of care between prisons and communities, carceral facility transfers and release from incarceration may disrupt continuity of care. In this study, we linked incarceration and TB databases to determine the effect of carceral movements on TB treatment outcomes in a Brazilian state. Our findings revealed that approximately one in three incarcerated individuals are transferred and/or released from the facility where they initiated treatment during the six-month TB treatment period. Both releases and transfers were associated with increased risk of unfavorable treatment outcomes, after adjusting for covariates. Individuals released less than two months into treatment had particularly low rates of successful treatment completion. These findings underscore the need for strategies to enhance continuity of care for incarcerated individuals with TB who are transferred between carceral facilities or released from incarceration during treatment.

Our findings provide novel insights into barriers to treatment completion among PDL in Brazil, identifying carceral movements as a major risk factor for unfavorable treatment outcomes in this population. Previous literature in the field has indicated that PDL have better TB treatment outcomes compared to the general population [7–12]. Although this may be true for a subset of patients who remain in the same facility during treatment, incarceration is dynamic, and facility transfers and releases are common. To date, studies in Brazil have been unable to test the effect of such movements on TB treatment outcomes due to the lack of detailed incarceration data in TB notifications databases. Our database linkage approach addresses these data limitations, enabling ascertainment of carceral movements during TB treatment and assessment of associated treatment outcomes.

Here, we find low TB treatment success rates among individuals who are transferred or released from incarceration during treatment, comparable to those of other highly vulnerable groups including people living with HIV, people who are unhoused, and people who use drugs [32–36]. Moreover, the treatment success rate among all PDL in our cohort, regardless of carceral movement group, was lower than in previous studies from Brazil which report treatment success rates exceeding 80% among PDL [7,10]. This discrepancy may stem from differences in the inclusion or classification of missing outcomes: while many prior studies exclude individuals without reported outcomes, we classify the lack of status update in the TB database as an unfavorable outcome. Addressing incomplete or delayed reporting in administrative databases is essential to improve monitoring and evaluation of TB treatment outcomes.

Our study has several limitations. Missingness within variables in the TB database (SINAN) prevented us from including several covariates in our analysis, such as smoking or drug use, which have been found to be associated with TB treatment outcomes [37–39]. Furthermore, there may have been residual confounding by other socioeconomic or incarceration-related covariates that were not available in our data. Our analysis may be subject to survivorship bias, as individuals in the transferred or released groups had to survive long enough to experience carceral movements. If survival is associated with treatment success, this could bias our results towards the null, leading us to underestimate the relative risk of unfavorable treatment outcomes associated with carceral movements. However, the low proportion of deaths in our cohort suggests minimal impact. Moreover, in our sensitivity analysis excluding individuals with non-TB deaths within six months of treatment initiation, the estimates of relative risk associated with carceral movements remained largely unchanged (Table S6, Additional File 1). Another limitation is that individuals categorized as released in our analysis may have previously experienced transfers that contributed to treatment disruptions. We did not differentiate between individuals released directly from incarceration to the community and those who experienced one or more transfers prior to release. We also do not differentiate between individuals released following the end of their sentence and those who escaped from incarceration. Additionally, we were unable to obtain treatment outcome data for individuals who moved outside the state of Mato Grosso do Sul during the follow-up period. Finally, the results of this study may not generalize to other states or countries.

Our findings highlight the urgent need for comprehensive strategies to improve continuity of care for individuals diagnosed with TB who are transitioning within and out of the carceral system. Ensuring timely transfer of medical records when individuals move to different carceral facilities is critical, especially for individuals transferred or released early during treatment. Referrals to community health care clinics, case management services, and discharge planning may play pivotal roles in facilitating continuity of care post-release [40–41]. In addition to improving system-level coordination, efforts to support individuals’ basic needs and social reintegration are essential. Individuals who are released from incarceration may face social instability, marginalization, and stigma, all of which can hinder care-seeking and access [42–45]. In the present study, we did not distinguish between people who were released versus those who escaped from incarceration, but the latter may experience heightened barriers to accessing healthcare. Further work is needed to understand these challenges and develop strategies to ensure all individuals, regardless of legal status or incarceration history, are supported to access healthcare services without fear of stigma or punishment.

## Conclusion

Our study found that both releases and transfers from carceral facilities were associated with increased risk of unfavorable treatment outcomes. Furthermore, individuals released earlier within their treatment regimen had higher loss to follow up rates. Our results highlight the need for strategies to ensure continuity of care for people with TB who are impacted by incarceration. These strategies can have broader implications for continuity of care after incarceration for health conditions beyond TB, a topic that remains poorly understood in Brazil and many other low- and middle-income countries [46–49].

## List of abbreviations

TB: Tuberculosis
PDL: Persons deprived of liberty
aRR: Adjusted relative risk
SINAN: Sistema de Informação de Agravos de Notificação
SIGO: Sistema Integrado de Gestão Operacional
SIM: Sistema de Informações Sobre Mortalidade
DR-TB: Drug-resistant Tuberculosis

## Additional Materials

File name: Additional file 1

File format: additional_file.pdf

Description: This file contains additional information on the methodology of our study as well as supplementary figures and tables.

## Declarations

### Ethics approval and consent to participate

The study was approved by the institutional review boards (IRBs) of Federal University of Mato Grosso do Sul (UFMS) (#7.060.054) and Stanford University (#50466).

### Consent for publication

Not applicable

### Availability of data and materials

This study utilized datasets provided by state agencies in Mato Grosso do Sul, Brazil, and we are not authorized to share them directly. The incarceration database was provided by Agência Estadual de Administração do Sistema Penitenciário (AGEPEN, https://www.agepen.ms.gov.br/). The tuberculosis and mortality databases were provided by the Secretaria de Estado de Saúde (https://www.saude.ms.gov.br/).

### Authors’ contributions

YM, YEL, JRA, and JC conceived and designed the study. EFL, MLDA, MCE and CG acquired the data. YM conducted the analysis and wrote the initial draft of the manuscript. JRA, YEL, JC, JB, and EFL provided critical input on analyses. YM and YEL contributed to writing subsequent drafts. All authors were involved in interpreting the data and critically revising the manuscript. All authors approved the final manuscript.

## Acknowledgements

Not applicable

