## Additional File 1 for "Tuberculosis treatment outcomes after transfer or release from incarceration: A retrospective cohort study from Brazil"

**Text S1 Data deduplication/linkage and cohort construction details**

*SIGO deduplication strategy details*

We conducted internal record linkage within SIGO using three key identifiers: name, mother's name, and an internal identification number (RGI). We used three different identifiers to mitigate potential challenges in matching entries due to missingness in mother's names, typos in names or mother's names, or inconsistencies in the recording of RGI numbers. Record linkage was performed in three stages:

1. RGI-Based Matching: Records hat had the same RGI were grouped together based on approximate string matching on names, with scores that exceeded specific thresholds.
2. Name-Based Matching Across RGIs: Records with the same names and mother's names but different RGIs were compared and consolidated based on defined thresholds.
3. Full Database Matching: All records in the database were compared against one another and consolidated based on defined thresholds.

To reduce the total number of string-matching comparisons, we only performed fuzzy string matching on pairs of records that met a low initial threshold of similarity. Specifically, we computed character-level 4-grams for all names which represent all contiguous 4-character length sequences within a name (eg. The name 'Miguel' yields the 4-gram sequence 'Migu, igue, guel'). We computed a 4-gram similarity score by calculating the proportion of shared substrings within the 4-grams of two names. If at least 70% of substrings in both of the names were shared, then we proceeded to perform fuzzy string matching.

All thresholds were determined through manual review of records with the same RGI. We identified records that belonged to the same individuals and set thresholds accordingly.

We utilized three algorithms for fuzzy string matching:

1. Levenshtein Distance: This metric was applied to compute similarity scores for names and mother's names.
2. Space Score: A modified Levenshtein Distance that is computed by obtaining the Levenshtein Distance between two names after removing all white spaces in the names. We incorporated this score into our matching process to address inconsistencies in the spacing of names in the SIGO database.

3. Partial Matching Score: This score is designed to handle missing portions of names such as a missing middle name within a record. This score tokenizes the shorter of two names using white space delimiters. It compares each token to all adjacent substrings of the same length in the longer name. The maximum Levenshtein distance for each token compared over all substrings is stored, and these scores are averaged to generate the final partial matching score.

The space score and partial matching score were only computed if the Levenshtein Distance between two names were sufficiently high.

*SINAN deduplication strategy details*

To deduplicate SINAN entries, we used the 4-gram approach and the string-matching functions previously used in the SIGO deduplication process. We additionally incorporated date of birth comparisons between records. We set thresholds after manually examining entries with identical names and mother's names but different dates of birth, as well as entries with identical dates of birth but differing names and mother's names.

*SIGO and SINAN database linkage approach*

To link entries between SIGO and SINAN, we compared names and mother's names using the same matching functions used in the deduplication processes of the databases. We similarly eliminated comparisons by utilizing 4-grams. We determined thresholds for linkage by examining the sensitivity and specificity over a range of values. To calculate sensitivity, we constructed a true positive set of entries in SINAN that we expected to match to entries in SIGO. This true positive set consisted of individuals who were recorded as being incarcerated (using the POP_LIBER and INSTITUCIO variables) in SINAN in 2012 and later, during which reporting of incarceration status improved in SINAN. To calculate specificity, we similarly constructed a true negative set of entries that should not match to SIGO records. This true negative set included individuals aged 12 and under in SINAN as of December 31$^{st}$ 2018 (the final date for which SIGO data is available in our study), as individuals aged 12 and under would not be incarcerated. Our matching process yielded a sensitivity of 90%, a specificity of 95%, and positive predictive value of 98%.

*SINAN and SIGO cohort filtering*

After matching records between SINAN and SIGO, we obtained 9,172 potential matches. Given imperfect specificity of our matching algorithm, we performed additional filtering to remove false positives. We excluded 27 matches where individuals had reported carceral movements in SIGO prior to their date of birth in SINAN. We further eliminated 90 matches of individuals who were minors at their date of TB notification. Our matching approach between SINAN and SIGO entries allowed for entries in either database to be matched more than once. In order to eliminate matches for individuals matched to multiple records, we selected the match pair with the highest matching score. Note, in the case where multiple SIGO records were matched to the same SINAN record, if the carceral movement history indicated that two records belong to the same individual (eg. identical origins and destinations for recorded movements as well as a sequence of dates that aligns for both records), we consolidated these matches. Following this additional filtering, we obtained a sample of 5,637 individuals matched across SINAN and SIGO. We then subset this sample to only include the 1,274 individuals who were first diagnosed with TB and initiated TB treatment while incarcerated between January 1st 2006 and June 30th 2018.

*SIGO, SINAN, and SIM database linkage*

To link entries from our SIGO-SINAN cohort with SIM, we applied the 4-gram comparison strategy used previously and compared names, mother's names, and dates of birth. For each entry in our cohort, matching was performed using the information within the SINAN database as it exhibited less missingness and fewer inconsistencies compared to SIGO.

We relied on deaths reported in both SINAN and SIM for recording outcomes in our cohort. We classified deaths as being TB-related and non-TB related. For records where at least one record indicated that the death was TB-related, we reported the death as TB-related. For discrepancies within the date of deaths, we used the earliest date in either system to address delays in reporting. All discrepancies in dates of death were of less than one month. At eight months following date of notification, six deaths were reported in SINAN that were not reported in SIM. 11 deaths were reported in SIM that were not recorded in SINAN. At two years following date of notification, nine deaths were reported in SINAN that were not present in SIM and 29 deaths were reported in SIM that were not present in SINAN.

*SIGO, SINAN, and SIM cohort filtering*

We retrieved 136 unique matches between SINAN and SIM. 114 of these matches had deaths within our study's observation period. We manually removed 4 false positive matches that included carceral movements or new TB cases reported after date of death in SIM.

*Group and Outcome Ascertainment*

Individuals are classified into three carceral movement categories based on their most recent movement prior to experiencing a terminal outcome within six months of their date of notification. Terminal outcomes are defined as events that determine treatment success within the observation period. For the purpose of our analysis, we assume TB treatment requires six months for completion. The observation periods and corresponding terminal outcomes are listed below:

**8-month observation period:**

1. Lost to follow up with no resumption of treatment within 2 months
2. Treatment Success
3. Death
4. DR-TB
5. Change of diagnosis/regimen

**2-year observation period:**

1. Treatment Success
2. Death
3. DR-TB
4. Change of diagnosis/regimen

### Table S1. Regression covariate missingness and imputation error

*Regression covariate missingness and imputation results*

| Variables | Missingness | Imputation Error |
|---|---|---|
| Alcohol use disorder | 24% | 25% |
| Age | <1% | 87 days |
| Mental Illness | 23% | 8% |

*Out-of-bag error is reported for each variable with missing values utilized in our regression analysis. For categorical variables, the imputation error is the proportion of false classifications. For continuous variables, the imputation error is the mean squared error. All regression covariates not included within this table, except for race and education, had 0% missingness. Due to high missingness for race and education, we did not utilize these imputed variables in our regression model and instead grouped missing values in the existing 'not reported' category within the data.*

### Table S2. Date of notification for incarceration cohort

*Distribution of Year of Notification*

| Year of Notification | Stationary (N=849) | Transfer (N=259) | Release (N=166) |
|---|---|---|---|
| **2018** | 76 (9.0%) | 32 (12.4%) | 17 (10.2%) |
| **2017** | 105 (12.4%) | 33 (12.7%) | 19 (11.4%) |
| **2016** | 129 (15.2%) | 25 (9.7%) | 18 (10.8%) |
| **2015** | 79 (9.3%) | 36 (13.9%) | 15 (9.0%) |
| **2014** | 85 (10.0%) | 20 (7.7%) | 13 (7.8%) |
| **2013** | 97 (11.4%) | 29 (11.2%) | 15 (9.0%) |
| **2012** | 83 (9.8%) | 31 (12.0%) | 16 (9.6%) |
| **2011** | 54 (6.4%) | 19 (7.3%) | 14 (8.4%) |
| **2010** | 43 (5.1%) | 11 (4.2%) | 4 (2.4%) |
| **2009** | 40 (4.7%) | 13 (5.0%) | 21 (12.7%) |
| **2008** | 38 (4.5%) | 7 (2.7%) | 9 (5.4%) |
| **2007** | 13 (1.5%) | 3 (1.2%) | 4 (2.4%) |
| **2006** | 7 (0.8%) | 0 (0.0%) | 1 (0.6%) |

### Table S3. Carceral facility units for incarceration cohort

*Distribution of individuals who were diagnosed and initiated treatment in each carceral facility.*

| Carceral Facility | Stationary (N=849) | Transfer (N=259) | Release (N=166) |
|---|---|---|---|
| **EPJFC** | 251 (29.6%) | 83 (32.0%) | 30 (18.1%) |
| **IPCG** | 105 (12.4%) | 46 (17.8%) | 14 (8.4%) |
| **PED** | 120 (14.1%) | 22 (8.5%) | 6 (3.6%) |
| **CPAIG** | 34 (4.0%) | 17 (6.6%) | 16 (9.6%) |
| **PSMN** | 32 (3.8%) | 7 (2.7%) | 7 (4.2%) |
| **EPMRSA-D** | 22 (2.6%) | 6 (2.3%) | 14 (8.4%) |
| **EPC** | 26 (3.1%) | 8 (3.1%) | 7 (4.2%) |
| **EPPAR** | 22 (2.6%) | 6 (2.3%) | 2 (1.2%) |
| **EPRACA-CG** | 14 (1.6%) | 3 (1.2%) | 13 (7.8%) |
| **PSM-TL** | 17 (2.0%) | 3 (1.2%) | 4 (2.4%) |
| **PDIB** | 16 (1.9%) | 6 (2.3%) | 2 (1.2%) |
| **CPA** | 6 (0.7%) | 8 (3.1%) | 7 (4.2%) |
| **EPAM** | 12 (1.4%) | 4 (1.5%) | 3 (1.8%) |
| **EPA** | 8 (0.9%) | 6 (2.3%) | 2 (1.2%) |
| **Other** | 164 (19.3%) | 34 (13.1%) | 39 (23.5%) |

*Carceral facilities within this table are the top 14 most populated within the cohort. All other facilities are grouped into 'other'.*

### Table S4. TB treatment outcomes two years following treatment initiation

*TB treatment outcomes 2 years post date of notification*

| Treatment Outcome | Stationary Group (N=849) | Transfer Group (N=259) | Release Group (N=166) |
|---|---|---|---|
| **Treatment success** | 626 (73.7%) | 168 (64.9%) | 90 (54.2%) |
| **No case status update** | 79 (9.3%) | 27 (10.4%) | 18 (10.8%) |
| **Referred to different health facility with no follow-up record** | 27 (3.2%) | 31 (12.0%) | 15 (9%) |
| **Treatment discontinuation with no follow-up record** | 60 (7.1%) | 23 (8.9%) | 34 (20.5%) |
| **Referred to different health facility/treatment discontinuation and resumed with no case status update** | 0 (0.0%) | 0 (0.0%) | 0 (0.0%) |
| **Death TB** | 13 (1.5%) | 1 (0.4%) | 2 (1.2%) |
| **Death non-TB** | 36 (4.2%) | 4 (1.5%) | 4 (2.4%) |
| **DR-TB** | 1 (0.1%) | 1 (0.4%) | 1 (0.6%) |
| **Change of diagnosis/regimen** | 7 (0.8%) | 4 (1.5%) | 2 (1.2%) |

*Carceral movement subgroups were determined at six months following treatment initiation and individuals may have experienced additional carceral movements before the end of the follow up period.*

### Table S5. Adjusted relative risk associated with unfavorable treatment outcomes 8 months post date of notification (extended table)

*Adjusted relative risk associated with unfavorable treatment outcomes (extended)*

| Characteristics | Categories | aRR | 95% CI |
|---|---|---|---|
| **Year of Notification** | 2017-2018 | Reference | Reference |
| | 2016 | 1.1 | 0.8-1.4 |
| | 2015 | 0.8 | 0.6-1.2 |
| | 2014 | 0.8 | 0.6-1.2 |
| | 2013 | 0.9 | 0.7-1.2 |
| | 2012 | 1.3 | 1.0-1.6 |
| | 2011 | 1.3 | 0.9-1.7 |
| | 2010 | 1.0 | 0.7-1.5 |
| | 2009 | 0.7 | 0.5-1.1 |
| | 2008 | 1.2 | 0.8-1.8 |
| | 2007 | 1.6 | 1.0-2.5 |

*Includes relative risks of additional variables for the regression model used in the primary analysis (Figure 2 and Figure 3).*

**Table S6. Adjusted relative risk associated with unfavorable treatment outcomes 8 months post date of notification (sensitivity analyses)**

*Adjusted relative risk associated with unfavorable treatment outcomes (sensitivity analyses)*

| Characteristics | Categories | Model 1 | | Model 2 | |
|---|---|---|---|---|---|
| | | **aRR** | **95% CI** | **aRR** | **95% CI** |
| **Incarceration Movement** | Stationary | Reference | Reference | Reference | Reference |
| | Transfer | 1.3 | 1.1-1.6*** | 1.5 | 1.2-1.8*** |
| | Release | 1.7 | 1.3-2.0*** | 1.7 | 1.4-2.1*** |
| **Facility Type** | Closed prison | Reference | Reference | Reference | Reference |
| | Semi-open prison | 1.1 | 0.7-1.9 | 1.6 | 1.3-2.0*** |
| | Police detention | 1.0 | 0.6-1.5 | 1.1 | 0.8-1.4 |

*Includes relative risks for variations of the regression model used in the primary analysis. Model 1 has the same structure as the primary regression model but additionally adjusts for individual carceral facility units. Model 2 has the same covariates as the primary model but excludes individuals who died from non-TB deaths within the first sixth months after treatment initiation. *** refers to .01 <p value <=.001. ** refers to 0.05 < p value <=.01. * refers to p value <=.05.*

**Table S7. Adjusted relative risk associated with unfavorable treatment outcomes 8 months post date of notification with released individuals stratified by timing of release**

*Adjusted relative risk associated with unfavorable treatment outcomes (with released group stratified by timing of release)*

| Characteristics | Categories | aRR | 95% CI |
|---|---|---|---|
| **Incarceration Movement** | Stationary | Reference | Reference |
| | Transfer | 1.4 | 1.2-1.7*** |
| | Release (<2 months post-notification) | 2.1 | 1.6-2.6*** |
| | Release (2+ months post-notification) | 1.3 | 1.0-1.8* |
| **Facility Type** | Closed prison | Reference | Reference |
| | Semi-open prison | 1.6 | 1.3-1.9*** |
| | Police detention | 1.1 | 0.9-1.4 |

*Includes relative risks for regression model stratifying individuals who were released into two groups: those who were released less than two months after treatment initiation and those released two or more months following treatment initiation.*** refers to .01 <p value <=.001. ** refers to 0.05 < p value <=.01. * refers to p value <=.05.*

**Figure S1 SIGO and SINAN matching criteria**

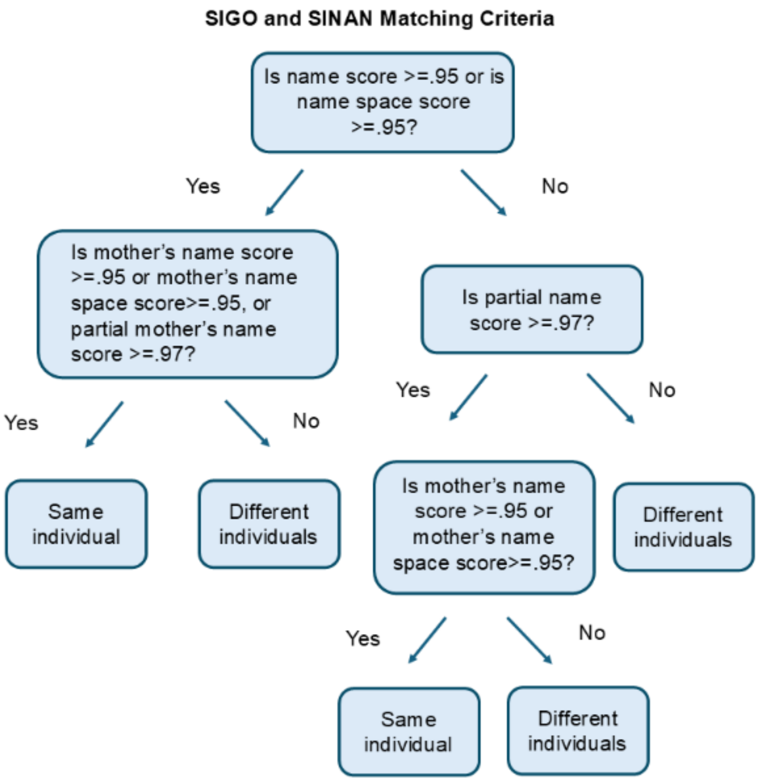


Decision tree depicting criteria used for identifying individuals between the SIGO and SINAN databases. String similarity was computed using functions described in Text S1.

**Figure S2 Carceral movements by facility type**

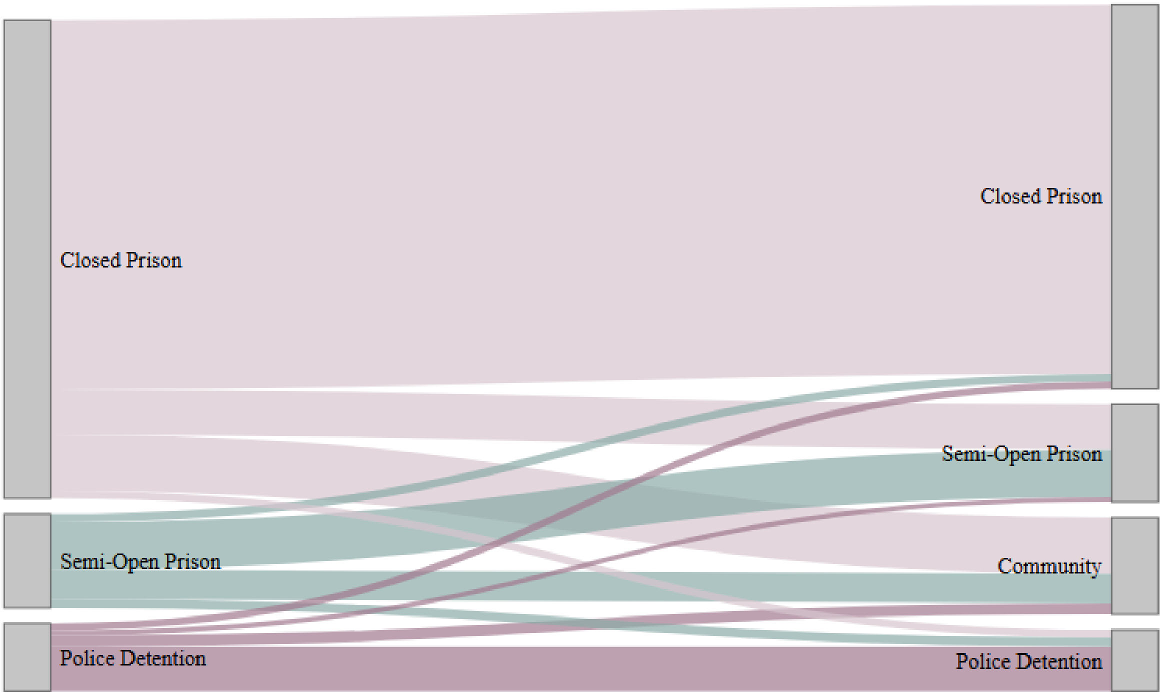


Sankey diagram showing carceral movements from the carceral facility location at the time of TB notification to the carceral facility location six months post-notification. Note that individuals may have experienced transfers between different locations of the same facility type (i.e., from one closed prison to another).

**Figure S3 Adjusted relative risks associated with unfavorable treatment outcomes within 8 months of notification (demographic and clinical variables)**

| Variable | aRR | 95% CI |
|---|---|---|
| **Sex** | | |
| Male | 1 | Reference |
| Female | 1.1 | 0.7 - 1.8 |
| **Age** | | |
| 18-29 | 1 | Reference |
| 30-44 | 1 | 0.9 - 1.2 |
| >45 | 1.1 | 0.8 - 1.4 |
| **Race** | | |
| White | 1 | Reference |
| Black | 1 | 0.8 - 1.4 |
| Asian | 1.2 | 0.7 - 2.2 |
| Brown | 1.1 | 0.9 - 1.3 |
| Indigenous | 0.6 | 0.3 - 1.3 |
| Not reported | 1 | 0.8 - 1.3 |
| **Education** | | |
| Did not complete high school | 1 | Reference |
| Completed high school/higher education | 0.5 | 0.3 - 0.9 * |
| Not reported | 1 | 0.8 - 1.2 |
| **Alcohol use disorder** | | |
| No | 1 | Reference |
| Yes | 1 | 0.8 - 1.3 |
| **Mental illness** | | |
| No | 1 | Reference |
| Yes | 0.7 | 0.4 - 1.3 |

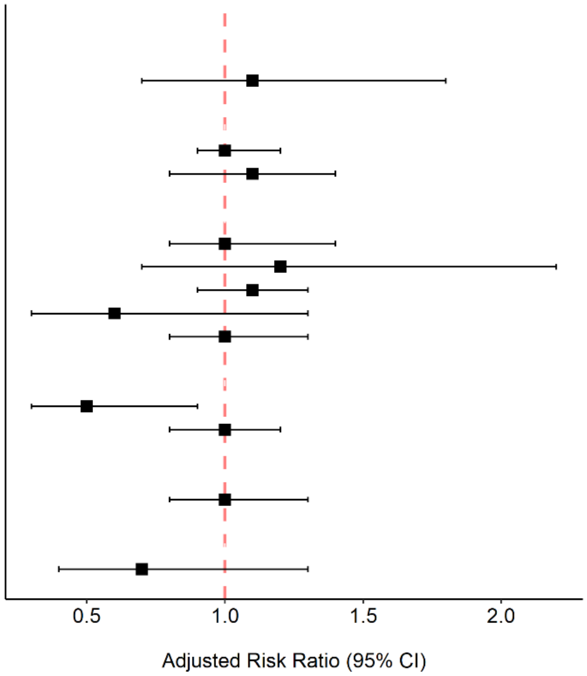


Adjusted relative risks of unfavorable treatment outcomes evaluated eight months after date of notification for demographic and clinical variables. Unfavorable outcomes refer to all outcomes other than treatment success.

**Figure S4 Distribution of time spent incarcerated from diagnosis to initial release**

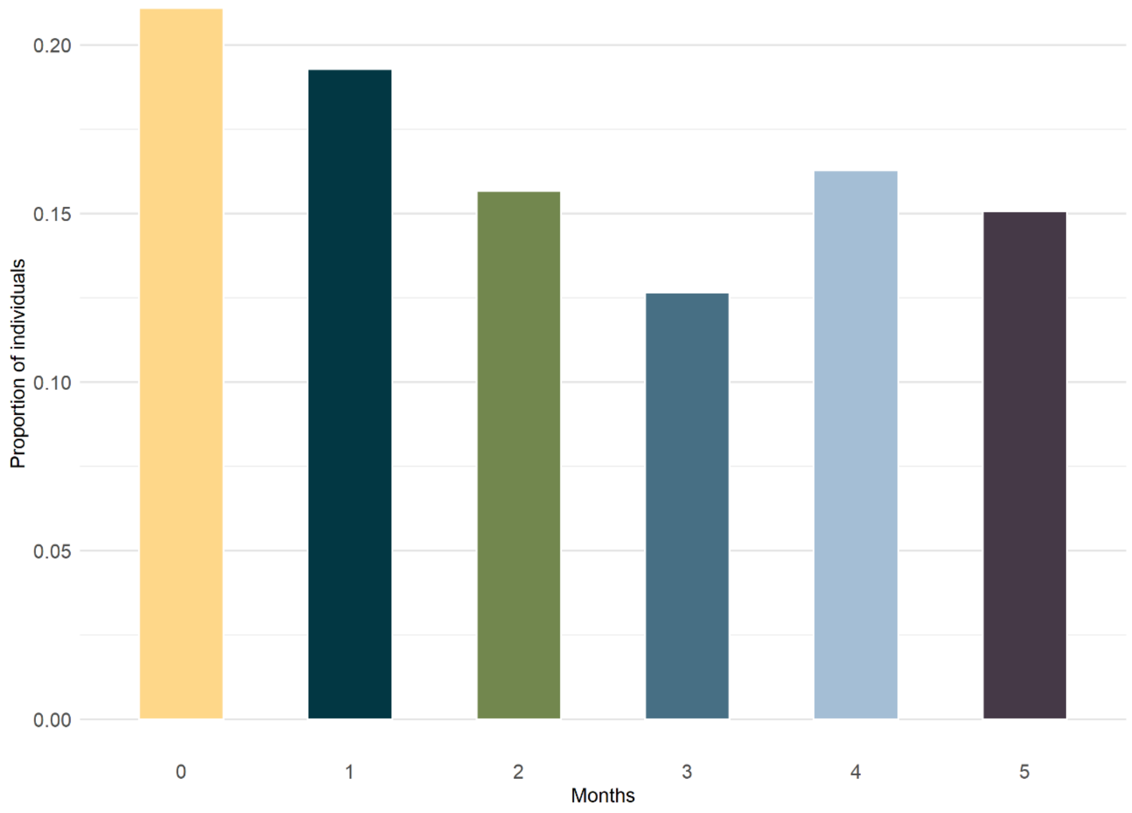


Bar chart depicting the time in months individuals spent incarcerated after date of notification until initial release. This figure is comprised of individuals within the ‘release’ carceral movement category. The distribution of time individuals spent incarcerated prior to initial transfer can be found in (Figure S5, Additional File 1).

**Figure S5 Distribution of time spent incarcerated from diagnosis to initial transfer**

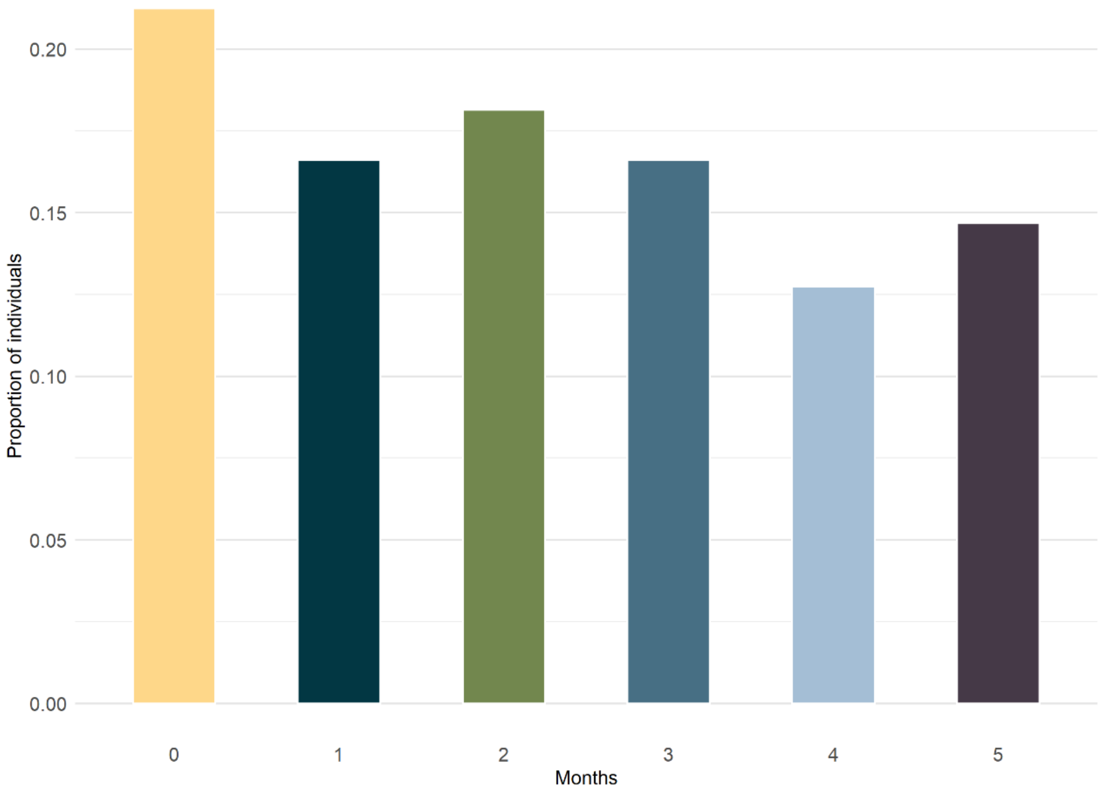


Bar chart depicting the time in months individuals spent incarcerated after date of notification until initial transfer. This figure is comprised of individuals within the 'transfer' carceral movement category.
